# Determination of COVID-19 parameters for an agent-based model: Easing or tightening control strategies

**DOI:** 10.1101/2020.06.20.20135186

**Authors:** Ali Najmi, Farshid Safarighouzhdi, Eric J. Miller, Raina MacIntyre, Taha H. Rashidi

## Abstract

Different agent-based models have been developed to estimate the spread progression of coronavirus disease 2019 (COVID-19) and to evaluate different control strategies to control outbreak of the infectious disease. While there are several estimation methods for the disease-specific parameters of COVID-19, they have been used for aggregate level models such as SIR and not for agent-based models. We propose a mathematical structure to determine parameter values of agent-based models considering the mutual effects of parameters. Then, we assess the extent to which different control strategies can intervene the transmission of COVID-19. Accordingly, we consider scenarios of easing social distancing restrictions, opening businesses, speed of enforcing control strategies and quarantining family members of isolated cases on the disease progression. We find the social distancing compliance level in the Sydney greater metropolitan area to be around 85%. Then we elaborate on consequences of easing the compliance level in the disease suppression. We also show that tight social distancing levels should be considered when the restrictions on businesses and activity participations are easing.

## 1. Introduction

Coronavirus disease 2019 (COVID-19) first emerged in Wuhan, China in December 2019 and is an ongoing pandemic caused by Severe Acute Respiratory Syndrome Coronavirus 2 (SARS-CoV-2). China implemented intense quarantine and social distancing and full lockdown of the cities in Hubei province on January 23^rd^ with the aim of controlling the pandemic, which has resulted into more than 81,000 reported cases so far (WHO Team, 2020). The disease has already gone outside Hubei, reaching to at least 26 countries in all parts of the globe by February. Control strategies of testing, tracing and lockdowns or other social distancing have been used in many other countries successfully, whilst countries which have delayed on lockdowns have had more severe epidemics. While the policies are effective and the pandemic has been largely controlled within China, the intense quarantine and full lockdown come with huge human and economic cost, which may not be acceptable in all countries. On the other hand, relaxing the restrictions can worsen the strain on the health care systems and threaten societies by resurgence of infection.

Enhanced surveillance and testing, case isolation, contact tracing and quarantine, social distancing, case isolation, household quarantine, teleworking, travel bans, closing businesses, and school closure are the most common strategies implemented worldwide for slowing down infection spread. While many of the strategies are currently in place in many countries, governments are looking for best policies for easing or lifting the control strategies. Thus, the extent to which restrictions can be lifted so that the disease remains under control and the economies do not suffer significant damage is a critical question.

While mathematical modelling of disease spread has a long history of providing solid foundations for understanding disease dynamics, the models are sometimes aggregated, with population heterogeneities ignored. These heterogeneities include zone population size and density, population age structure, age-specific mixing, the size and composition of households, and, critically, travel and activity participation patterns which may have important impacts on epidemic dynamics and on the effectiveness of possible interventions (Grefenstette et al., 2013). Recently, the development of disaggregated models in infectious disease epidemiology has received considerable attention owing to their capability to capture the dynamics of disease spread combined with the heterogeneous mixing and social networks of agents. While all the developed agent-based modes can capture complex interactions between agents and to some extent their decisions, they cannot consider the interactions of agents within a household and therefore, the interconnection between the travel decisions and activity participation of different household members. For example, teleworking may shift agents to participate in other activities or school closure may affect the activity patterns of all the household members. Building on the preliminary version of the SydneyGMA model, which is an activity-based model developed for the Sydney greater metropolitan area (GMA, see Figure A1), this paper focus on proposing a an agent-based disease transmission model estimation that accounts for interactions between people.

Furthermore, the virologic and epidemiologic characteristics of SARS-CoV-2, including transmissibility and mortality, are not yet fully known. Despite a surge of efforts to estimate the disease spread parameters, these parameter estimates typically show considerable variation from one study to another. They also are only applicable for aggregated models such as SIR-based models. To the best of our knowledge, there are no guidelines or research on the parameter estimation of pandemic diseases modelling in agent-based models, mainly due to the complexity of agent-based models and the existence of many interactive parameters. This paper contributes to the determination of COVID-19-specific parameters useful for agent-based modelling of disease spread. Further, of the few prior attempts to calibrate parameters (Chang et al., 2020), these efforts have been *unstructured* in that the interconnections among the parameters on the pandemic effects are not considered. Unstructured calibration refers to the sequential adjustments of parameters in a relatively ad hoc and non-systematic way. Although an unstructured calibration approach may reproduce observed statistics, the approach can be problematic for many reasons, including the failure to consider interactions among parameters, and excessive focus on reproducing observed statistics, at the possible sacrifice of model system validity. In this paper we use *response surface methodology* (RSM) to efficiently calibrate the model while considering the interactions of their constituent parameters. By optimally calibrating parameters, their unbiased impacts on disease spread can be captured. Given the observed statistics of the Sydney GMA, including the number of cases and public transport (PT) usage after lockdown, we calibrate the parameters for an agent-based model for the Sydney GMA. It is noteworthy to say that the transport activity-based model of Sydney includes the actual transport network with models reflecting the overall travelling behaviour of people in an urban metropolitan area.

After calibration of the transmission model parameters, we use the model to explore several scenarios examining the influences of easing social distancing restrictions, opening up businesses, timing of control strategies implementation, and quarantining family members of isolated cases to intervene the disease progression. The intent is to provide guidance to public health agencies worldwide as they consider easing of restrictions.

## 2. Model description and calibration

This section briefly explains the activity-based model used to model the pandemic spread and then, the methodology for model parameter calibration is introduced.

### 2.1 SydneyGMA model

The agent-based disease transmission model in this paper is built on an activity-based model developed for the Sydney GMA, called SydneyGMA model, which has several properties that are valuable for analysing the effectiveness of COVID-19 control strategies. Firstly, SydneyGMA uses the Travel/Activity Scheduler for Household Agents (TASHA), an operational, state-of-the-art model of daily travel and out-of-home activity participation that considers both individual activities as well as joint household activities, along with a full range of within-household interactions (Miller and Roorda, 2003; Miller et al., 2005; Roorda and Miller, 2006; Roorda et al., 2008, 2009; TMG, 2015). In addition to Sydney, TASHA has been applied in Toronto, Canada (where it is the operational model for Toronto transportation planning agencies), Helsinki, Finland, and Temuco, Chile. All parameters of the Toronto model are transferred to the Sydney model. Consequently, in the case of school closures or widespread working from home, the activities of households will be realistically rescheduled, factoring in the extra time derived from removing school- and work-related activities from the household’s regular schedule. Secondly, mode choice is computed for each household individually, and interactions between household members using their vehicle on individual or joint trips are captured, as well their usage of other modes of travel, notably transit. Thirdly, the model “assigns” transit (PT) trips to explicit paths through the transit network, enabling different components of transit trips (including in-vehicle, walking to/from transit, and waiting and transferring) to be estimated and considered as potential situations for disease spread. Therefore, utilising the SydneyGMA augments the disease spread modelling by accounting for potential locations of disease spread and more accurately modelling interactions among household members as a result of adjustments to their daily activities. The limitation of SydneyGMA is brought in Appendix B)

### 2.2 Disease spread parameter determination

The disease spread model, explained in Appendix C, iteratively interacts with SydneyGMA model once par day and scrutinises the itinerary of each agent in the system. Accordingly, it updates the disease state of each agent. In particular, the changes each day between agents’ disease states affect their travel behaviour and activity participation (and their family members itineraries) in subsequent days of the simulation.

There are several factors that affect the movement rates (probabilities) among the different disease states. The factors can be categorised into 1) travel behaviour-specific parameters, 2) disease-specific parameters, and 3) policy-specific parameters. The travel behaviour-specific parameters affect out-of-home activity participation rates, destination choices, travel mode choices, the start time, location and duration of out-of-home activity episodes, and contact number for activity type. Except for the contact number, the other parameters are transferred from the original TASHA model and adjusted for the Sydney context and integrated to the transport network of Sydney.

The disease-specific parameters include incubation period, average time required for an infected agent to recover, and the probabilities of: becoming infected (per contacted person), transitioning from infectious to quarantined (per day), infected agents dying (per day), and transitioning from quarantined to recovered (per day). In Appendix D, we describe the parameter calibration procedure used to determine the parameters for the agent-based disease spread models and present the resulting calibrated parameters in Table C1. The parameter calibration procedure is based on previously published work by Najmi et al. (2019b).

The strategy-specific parameters determine the policies that might be applied by policymakers and authorities to slow down disease spread. These include, but are not limited to, the enforcement of business closings, teleworking, and, if applicable, easing the restrictions on businesses; school closures and re-openings; infected case isolation; quarantining of family members; social distancing; and the dates when the restrictions are in place. Of these, variations in school closure strategy have not been considered in this paper due to the huge uncertainty that exists with respect to the impact of the virus on children. Another strategy-specific parameter is the change of trip generation rates, which is usually ignored in conventional disease spreading models.

## 3. Control strategies

We evaluate several different control strategies, namely: home quarantine of family members of the traced infected cases, social distancing, travel load reduction, and the date when the control strategies are imposed. Different scenarios are run to explore these control strategies and the dates when they are implemented. However, we do not explore the impact of case isolation (CI) and school closure (SC) in this paper. CI and SC strategies are set to our best estimate of current values for the Sydney GMA and are held constant across all experiments. We assume that CI is implemented from the start day of the epidemic, as has been the case in Australia and most other countries. The SC strategy comes into effect in the analysis in the week starting 23 March 2020. Early in this week, the schools were still open, but it was up to parents to decide whether to send their children to school or not. Thus, SC is considered to remove schools and universities from the list of activities for a majority of students. We assume that universities are partially open and 10% of university students continue to travel to universities in this scenario. Obviously, the SC affects the daily travel itinerary of the students and their family members. Studies have estimated that SC requires around 15% of the workforce to take time off work to care for children, which is associated with considerable costs (Scott, 2020). This changes in the activity participation is captured by SydneyGMA.

Scenario assumptions for each of the control strategies examined are briefly described in each of the following sub-sections.

### 3.1 Quarantined family members (QF)

QF is a common strategy to control pandemics. While different levels of quarantine strategies are implemented worldwide, we only investigate the existence or the lack of this strategy. In the case of existence, we assume that the strategy is implemented from the day of finding the first case in New South Wales (NSW), on 22 January 2020. Following identification of a symptomatic case in a household, all household members remain at home for 14 days.

### 3.2 Social distancing (SD)

SD is a key parameter in disease transmission models and affects the rate at which sick people infect susceptible people. We impose SD in our model by the adjustment of all non-household contacts (referred to as compliance level) while the intra-household contacts are kept unchanged which is in line with the pandemic studies of Chang et al. (2020) and Ferguson et al. (2020). So, the SD compliance levels may vary from zero-SD – no compliance-to full lockdown-full compliance, with a rate at which the contact rates are affected following the SD control strategy. This strategy came into effect in NSW on 31 March 2020.

### 3.3 Travel load (TL)

TL is used to reduce non-essential trips, including leisure, sport, and religious activities. Also, it includes the reduction in trips due to teleworking, layoffs, and quitting a job. Similar to SD, we define different levels and investigate the influence of enforcement to eliminate unnecessary trips. We consider the TL level in Sydney GMA in April 2020 as the extreme level in our investigations and explore the influences of easing the restrictions. Despite some rare cases, as in Wuhan, where the TL levels have approached 0%, in many other countries, the enforcement of the severe TL restrictions is impossible. The TL strategy comes into effect within the analysis starting from 23 March 2020.

### 3.4 Date of lockdown (DL)

The date when the control strategies are implemented is a controversial decision for authorities. This is a difficult decision for governments, as it has detrimental effects on economies and, in the worst case, might result in economic collapse.

It should be noted that an important effect of lockdowns is on travel behaviour, and, as a result, on urban travel demand. There is no current data that provide information about the changes in travel decisions of agents after lockdown. Thus, we need to make some assumptions, the most important of which is the travel volume after lockdown. As there is no reliable data on the generated trips after lockdown in Sydney GMA (in April 2020) compared to before, we assume 50% reductions in the total number of trips. However, according to Transport for NSW (Transport for NSW, 2020), the PT usage reduced by 79% after lockdown. So, the change in the PT usage is a piece of reliable information we used and adjusted the utility of PT mode in SydneyGMA to fit the simulated ratio to the observed statistic.

The next section explores the effects of implementing and relaxing each of these control strategies.

## 4. Runs and results

As the system is probabilistic, starting with very small number of infected cases (e.g. one or two cases) may substantially affect the simulation results, depending on whether the model quarantine them sooner or later. Thus, we use an initial set of four infected cases in the population. Because there have been four active cases in Sydney GMA in 28 February 2020, this date is selected as the starting point of experiments. Numerous simulation runs of the combined SydneyGMA and calibrated agent-based disease spread model were run and the simulation results are presented and discussed in the following subsections.

### 4.1 Base case

The base case scenario is equivalent to the settings that reproduce the observed statistics; thus, it is the output of the calibration model. Figure 1 shows the base case scenario obtained from the simulation of the ongoing spread of COVID-19 and reproduces the disease spread progression in SydneyGMA. In the scenario, all the control strategies are in place as in reality in NSW. The SD compliance level and TL level strategies after lockdown are determined and considered at 85.9%, called the *base* SD compliance level, and 50%, called the *base* TL level, respectively (see Appendix D). Figure 1 (A) and (B) reveal the high performance of our calibrated disease transmission model in reproducing the observed infected cases. As a result of the restrictions implemented by the Australian government in the last week of March 2020, the reinfection rate drops sharply, and the epidemic almost dies out. Figure 1(C) shows the simulation result of running the model in the base scenario. This figure distinguishes between the isolated (but not necessarily infectious) and non-isolated cases. So, the model estimates that about half of the persons in quarantined state are the family members that are not actually infected. In reality, while the family members of infected cases are quarantined, their infection to the disease has not yet been determined.

**Figure 1:**
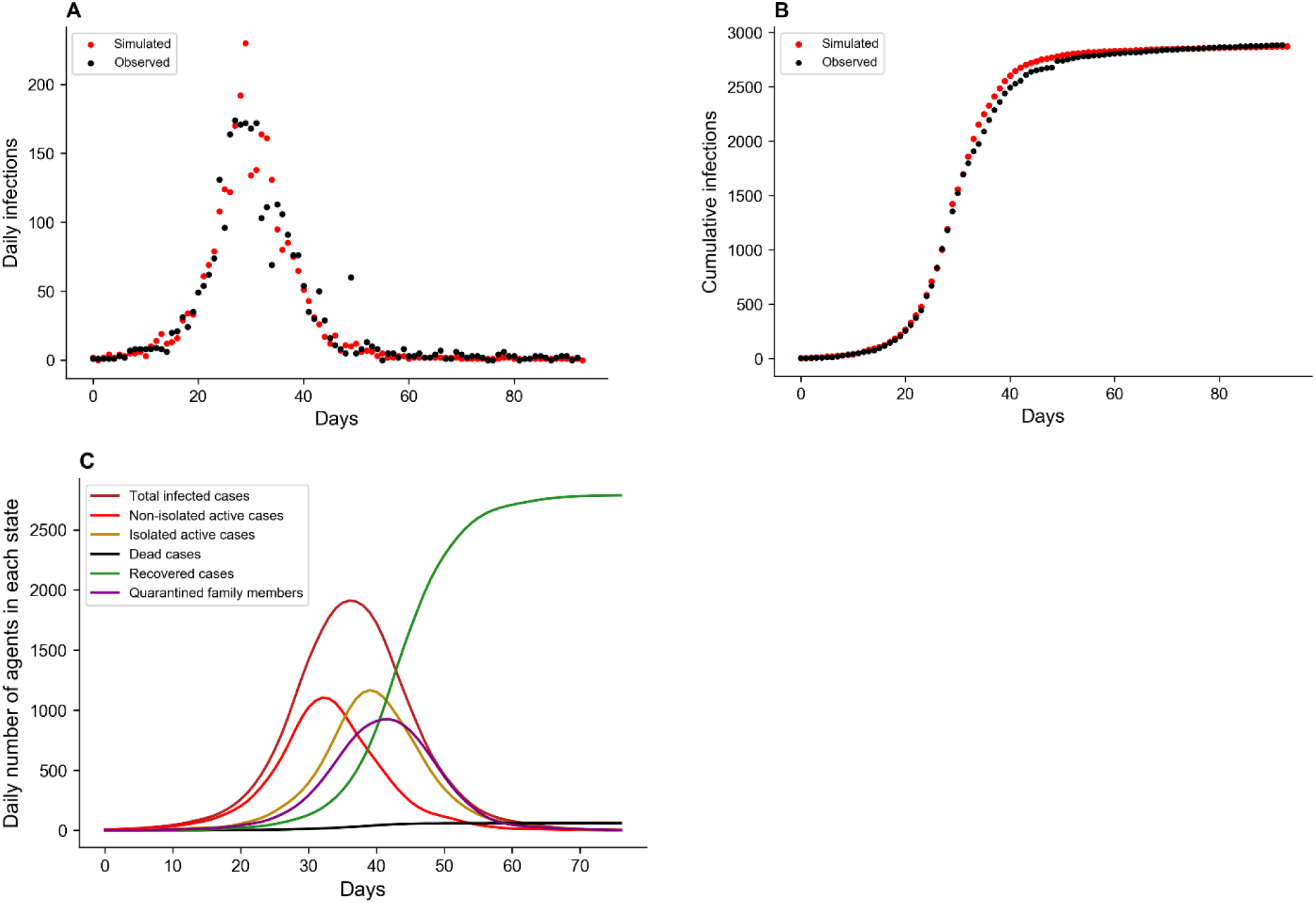
Power of the calibrated SydneyGMA -based disease spreading model in reproducing the daily number of cases (A), the cumulative number of cases (B) and the number of cases at each state of the pandemic modelling (C) in the base-case scenario.

### 4.2 Various SD Compliances

In model calibration, we found the base SD compliance level after shutdown in the Sydney GMA. However, the exploration of easing the compliance level on the disease distribution allows policymakers to identify the minimum compliance levels for which the disease might be controlled. Figure 2 shows the simulation results of the social distancing strategies, coupled with QF and base TL level, across different compliance levels. We do not consider the SD compliance level of 100% as it is almost impossible to achieve. The figure reveals that compliance levels of less than 70% do not show enough strength to suppress the disease within 3 months. At these compliance levels, the number of emerging new cases is higher than the potential of the health system to find and isolate the infected cases. While the SD base compliance level could eliminate the disease, or hold it close to zero cases, in about 2 months, the lower SD compliance levels of 80% and 70% could control the disease with a delay of 14 and 28 days respectively. Reducing the SD compliance by 15.9%, from 85.9% to 70%, increases the cumulative number of cases by 59%. Still, this is much better than the scenario in which there is 50% or less SD compliance level in place.

**Figure 2:**
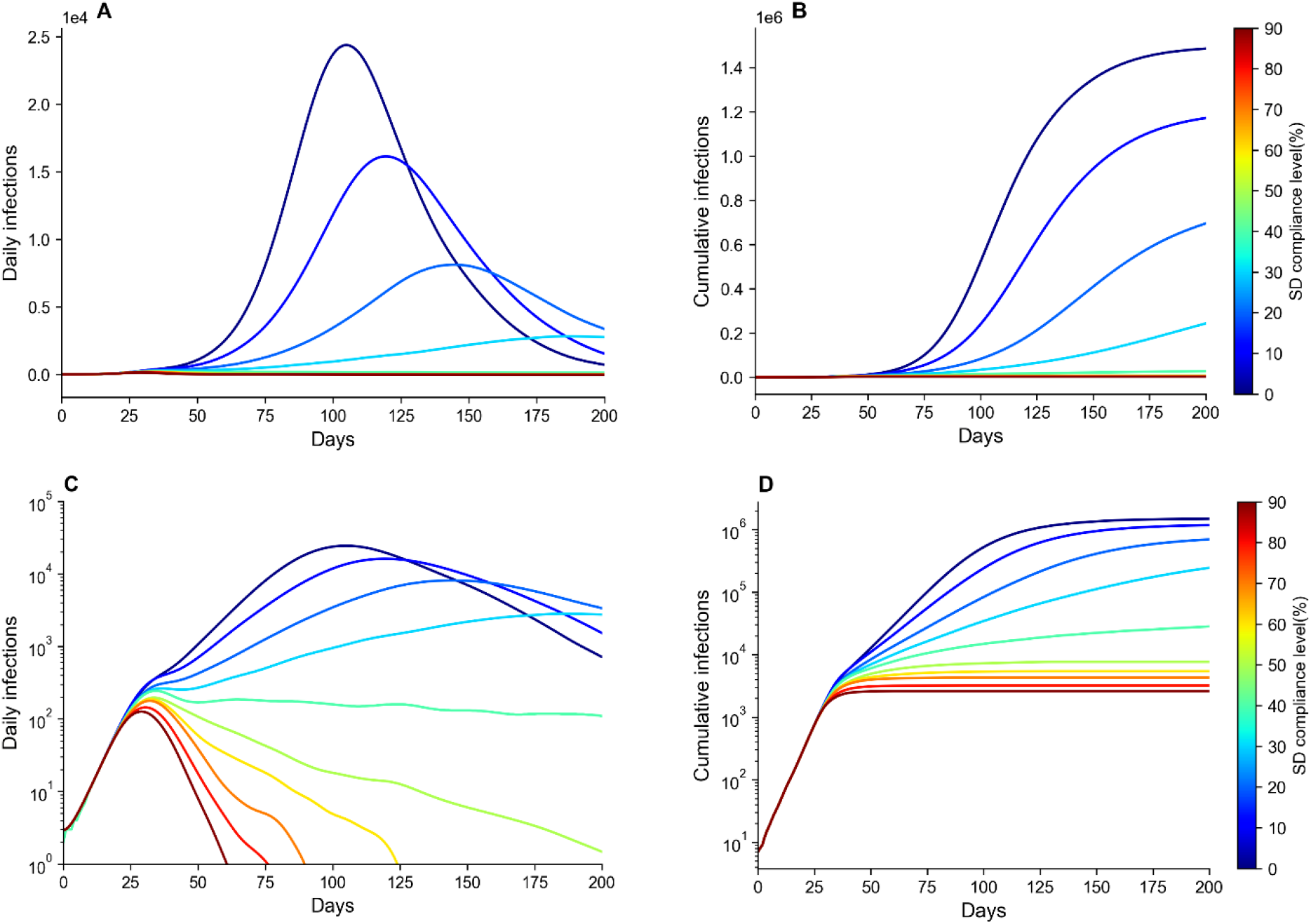
A comparison of different SD compliance levels. The settings for other control strategies are the same as in the base scenario. (a) daily number of cases (linear), (b) cumulative cases (linear), (c) daily number of cases (logarithmic), and (d) cumulative cases (logarithmic). Note: Responding to the skewness of large values, (A) and (B) are plotted in logarithmic scale in (C) and (D).

The compliance levels between 50% and 60% are still effective in reducing the infected cases (at base TL level), but they do not suppress the disease in a short period of time. Thus, control of the disease with these SD levels required a longer time period. In these cases, the resurgence of disease spreading is probable. The SD compliances levels of less than 50% are not strong enough, for any duration, to suppress the disease.

### 4.3 Speed of implementation of lockdown

We evaluate the timing of the implementation of lockdown in Sydney GMA. Figure 3 explores the scenarios where all the control strategy settings are the same as in the base case scenario, but they are enforced either 3 or 7 days earlier or later than the actual introduction date. This figure reveals the impact of selecting an appropriate time to apply the control strategies. Left unchecked, the spread of the disease grows exponentially such that in the first three weeks the number of infected agents is small, and the situation does not seem dramatic. Then, the values change rapidly. Earlier enforcement could lead to 96% and 16% fewer cases for the scenarios with the lockdown implemented 7 and 3 days sooner, respectively. The delays of 3 and 7 days, on the other hand, could lead to, respectively, 25.6% and 686% increases in the number of cases. A week’s delay not only increases the pressure on the health system considerably but also requires an approximately 30-day longer suppression period.

**Figure 3:**
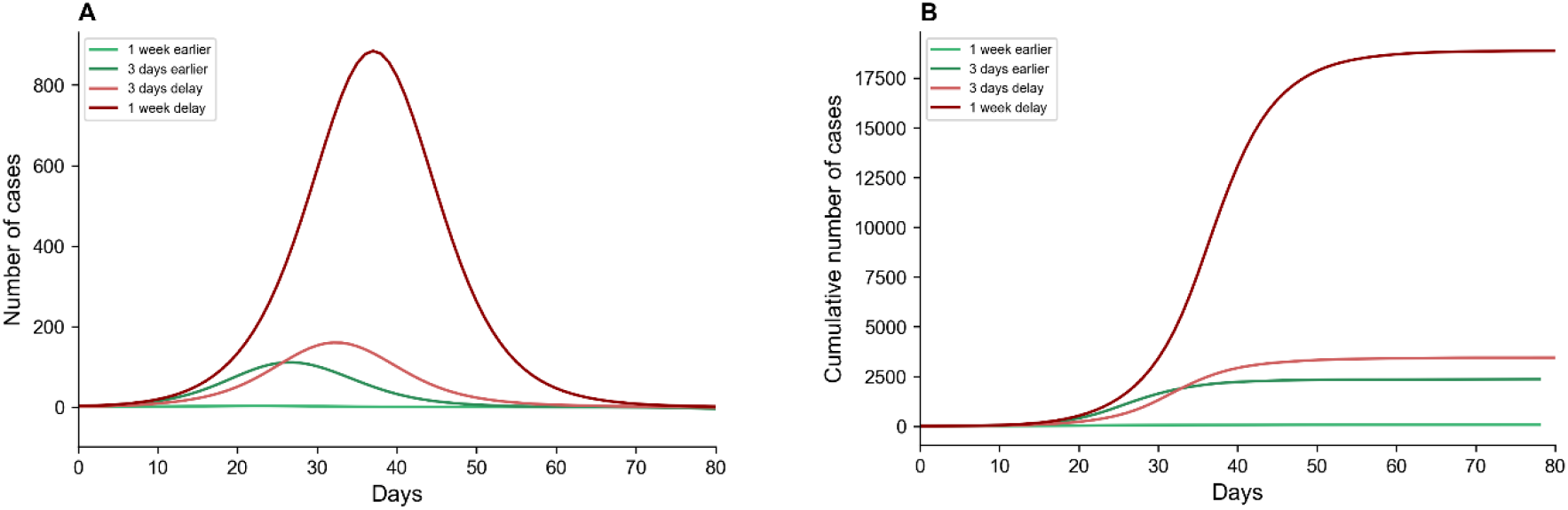
A comparison between the influence of implementing the lockdown earlier (in greenish) and later (in reddish) while all the strategies are in place as in the base case scenario. (A) Daily number of cases, and (B) Cumulative number of cases.

### 4.4 Opening businesses

In the base scenario, we defined the base SD compliance and base TL levels. Furthermore, in Figure 2, we show that compliance levels over 60% suppress the disease in a reasonable time (at the base TL level). Suppose that the generated trips increase by easing the restrictions on businesses to open again, but all other in-place strategies are still in effect. To examine this case, we run the model with different TL levels across both the base and 60% SD compliance levels to investigate the interaction effect of the SD and TL control strategies. The results of running the scenarios are presented in Figure 4. The figure also explores the importance of the QF control strategy in controlling the disease spread.

**Figure 4:**
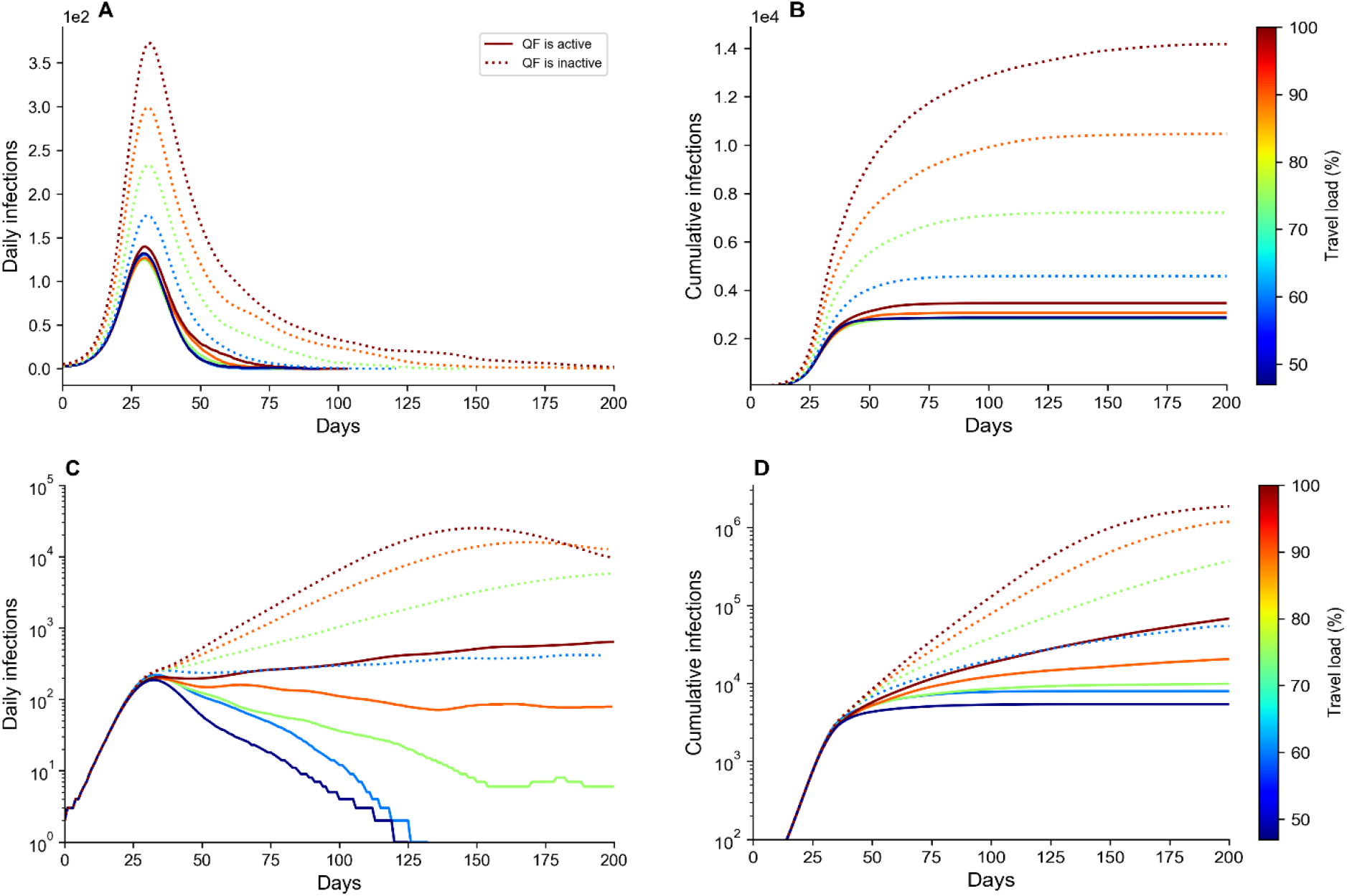
A comparison of different travel load and its interaction with home quarantine strategy at two social distance compliance levels of 85.9% and 60%. (A) daily number of cases (SD compliance levels is 85.9%), (B) cumulative cases daily number of cases (SD compliance levels is 85.9%), (C) daily number of cases (SD compliance levels is 60%), and (D) cumulative cases daily number of cases (SD compliance levels is 60%). Note: Responding to the skewness of large values, (C) and (D) are plotted in logarithmic scale.

Having the QF strategy in place throughout the period, the base SD compliance is very successful in controlling the disease spread progression in a short period of time for all the TL levels. However, while the SD compliance level of 60% is successful in suppressing the base travel load, the result is not satisfactory for travel loads of 80% and over. This reveals that even slightly easing the social distance controls while the travel demand is close to the pre-COVID-19 travel demand level, will be ineffective.

The figure also shows that relaxing the QF control strategy significantly increases the disease suppressing period, even if a high compliance level of social distancing is in place. Further, relaxing the QF multiplies the number of patients. It also remarkably increases the magnitude of the daily infections, especially when coupled with a low SD compliance level. Thus, the QF strategy has significant interaction effects on both travel load and SD compliance level, such that ignoring the QF strategy multiplies the daily infection rate and infected cases.

## 5. Conclusion

Australia has achieved excellent control of COVID-19, with very few new cases by June 2020. Travel bans remain in place and have been very successful in averting a much larger epidemic (Costantino et al., 2020). NSW had the largest number of cases, and the greatest challenges in disease control. As the country and state of NSW begins to open up again, on a backdrop of low disease incidence, mitigating resurgence of COVID-19 and maintaining the hard-won gains is critical. We show that the likely compliance with social distancing was 85.9% during the period of lockdown, and that reduction in compliance can result in disease resurgence. As society re-opens, enhanced surveillance and testing for COVID-19 is essential, and at the first signal of resurgence, lockdown should be implemented without delay. We show that a delay of even 1 week can be costly. A return to normal travel and use of public transport in Sydney GMA will result in a risk of resurgence, but can be mitigated. The use of face masks may also be key to safer resumption of travel within Sydney (Stutt et al., 2020).

## Data Availability

All data which we have addressed to is available.

## Appendix A

The study area is shown in Figure A1.

**Figure A1:**
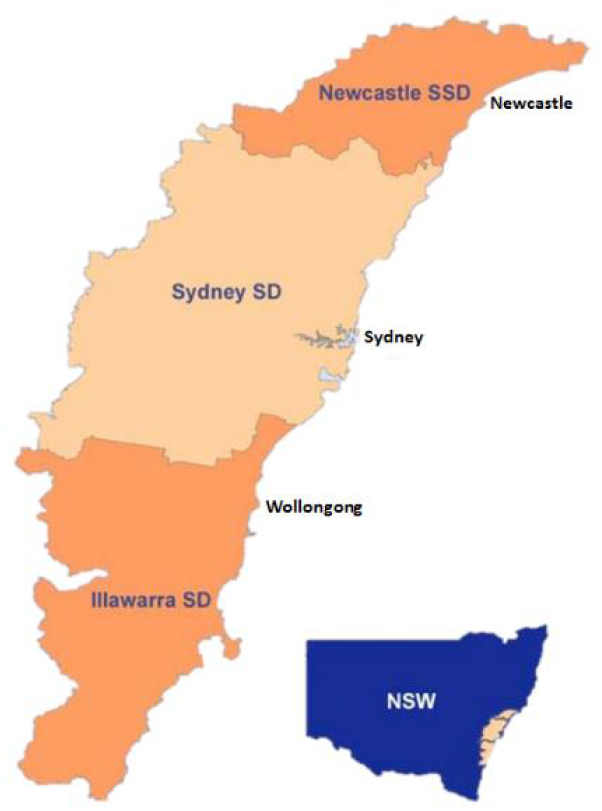
Sydney greater metropolitan area map

## Appendix B

### SydneyGMA limitations

Similar to every other model, SydneyGMA has several limitations. First, SydneyGMA is developed by transferring GTAModel V4.1, which is originally developed for Greater Toronto-Hamilton Area (GTHA). Transferring is a common approach in developing activity based models because there is no immediate need for re-estimation of parameters (Bowman et al., 2014; Castiglione et al., 2014). While the socioeconomic data, network datafiles and procedures for Sydney has been used for the development of SydneyGMA, there are still some parameters that need to be estimated and replaced. As the paper objective is to investigate different intervention policies comparatively, we believe that being at this stage of transferring does not affect our scenario analysis remarkably. Second, SydneyGMA incorporates all the travel and out-of-home activity participation decisions and household profiles of each agent, it is not able to explicitly model out-of-home interactions among agents at a micro “fact-to-face” level. While the model generates the activity type and the zone where an agent participates in an out-of-home activity, it does not provide detailed information about other agents who have been in contact with this agent at the specific activity location (e.g., a store) at the specific time of the agent’s visit to this location. So, we had randomly pick interacting agents from those who were in the same zone and activity level and overlap at the attendance time. The same assumption has been used for the PT mode. Third, the SydneyGMA does not provide the specific PT vehicle that is used by each agent. Thus, we select interacting agents from those that use the PT mode to get to the same destination and activity type with approaching times that are close to each other. Assumptions made about the transport model do not affect the conclusions drawn from the relative analysis conducted in section 4, as the intention of the paper is not to provide accurate forecasts, instead it is aimed to assess the relative costs and benefits of different conferment policies.

## Appendix C

### Proposed disease transmission model

The proposed disease transmission model is built on the SydneyGMA activity/travel agent-based microsimulation model. It stochastically models disease transmission due to inter-agent interactions while travelling and engaging in out-of-home activities, as well as within-household interactions. It sensitive to a wide range of disease control strategies affecting these interactions Seven classes of agents are included in the transmission model: 1) Susceptible agents (S), who are not in contact with infectious agents and are subject to be infected, 2) Exposed agents, who are infected agents but in the incubation period (latent) of the disease, 3) Infectious agents, who are contagious, 4) Quarantined agents, who are infected agents quarantined by health care authorities, 5) Quarantined family members, who are the family members of at least one infected and quarantined agent, 6) Dead agents, who are the infected persons, either in quarantine or not, that have died, and 7) Recovered agents, who are infected persons, either in quarantine or not, who have recovered.

Figure B1 a presents flow chart which explains in detail the sequence of the states in the proposed model. Firstly, all agents, except those who are infectious, are susceptible. The susceptible agents may come into contact with contagious agents and then may acquire the infection probabilistically and move to the exposed state. Exposed agents remain non-contagious for a given incubation period. In our simulation, the incubation period is a parameter that should be calibrated. At the end of the incubation period agents will become contagious and move into the infectious class. Infectious agents, both symptomatic and asymptomatic cases, become quarantined by health care authorities probabilistically. Thereby, they move into the quarantined class. Obviously, non-quarantined agents are the main source of disease spread in the system. Upon discovery of an infected agent, not only does the health care authority quarantine them, but they also quarantine their family members as they have a high chance of being exposed to the disease and may be in their incubation period. The quarantined family members who are infected will move into the Quarantine class. Quarantined agents may respond to treatment and recover, and move to recovered class, or die, and move to the dead class. Albeit, for COVID-19, infected but non-quarantined agents have also a small chance to die and move to the dead class, but we ignore this possibility in this paper. On recovery from infection, individuals are assumed to be immune to re-infection. All the infected agents, either quarantined or not, will be recovered after 14 days which is the typical recovery period of COVID-19.

**Figure B1:**
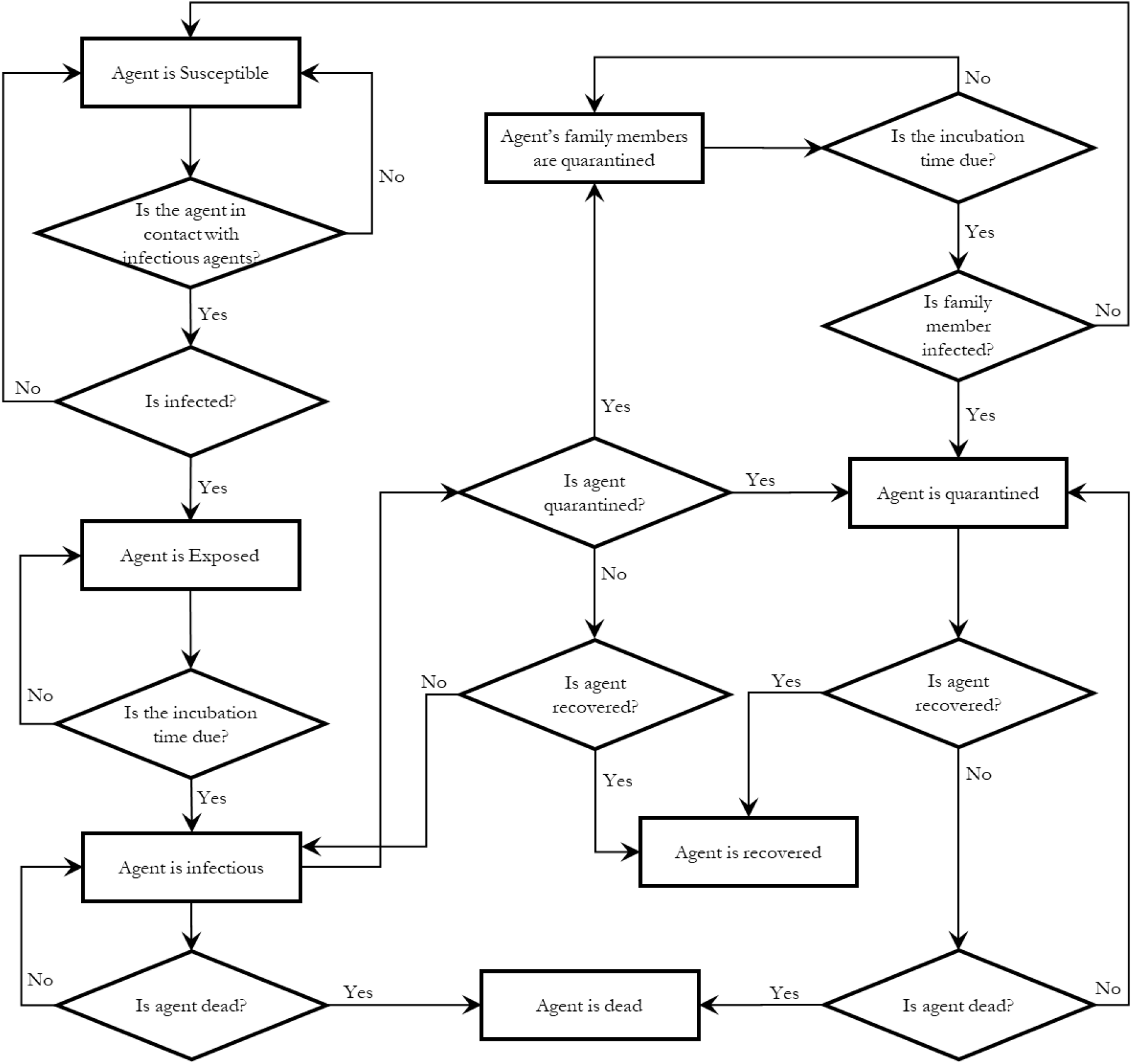
Flow chart of the proposed model

## Appendix D

### Model calibration approach

Parameters of disease transmission models including contact rate and infection rate are interconnected and their influences are complementary. The agent-based disease transmission models (including disease spreading models and ABMs) are highly non-convex, so a reliable mathematical model between the parameters (called dependent factors) and their effects (called responses of interest) are not available. We use a response surface methodology (RSM) to calibrate the disease transmission model. RSM consists of a set of mathematical and statistical techniques used to develop, improve and optimise processes in which a response of interest is influenced by several factors, with the eventual objective of optimising the response (Box and Draper 2007). Therefore, RSM quantifies the functional relationship between a response of interest, *y*, and the explanatory factors, *x* (See Eq. 1). This mathematical representation may correspond to different orders of polynomial functions (Najmi et al., 2019b). In this paper, we choose the order of polynomial corner points of each equation based on a fitness function value for the estimated equations. It should be emphasised that the interaction of the parameters can also be estimated mathematically. For a complete review of RSM techniques, the reader is referred to Khuri and Mukhopadhyay (2010).

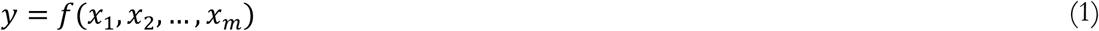

RSM shows how the parameters *x* affect outputs *y*, and allows determination of the independent parameters that optimise the output for calibration purposes. To conduct an RSM analysis, we use a central composite design (CCD) to design the experiments and analyse the results to obtain the optimal parameter values. CCD was firstly introduced in Box and Wilson (1951), as the design matrix because it allows reliable identification of first-order interactions between parameters while providing a second-order polynomial model to predict their optimum levels (Myers et al., 2009). The methodology is shown in Figure C1 for three factors. CCD uses three groups of corner points, centre points, and axial points to fit the functional relationship in Eq. 1. Corner points represent the factorial design points and are coded by ±1. In a centre point, coded as zero, the value of each factor is the median of the values used in the factorial portion. For axial points, all parameters are held constant at zero, except for one with the value of +α or – α, as a *design parameter* (Marget 2015). For more information about different experimental designs and the required number of experiments to be run, the reader is referred to Myers et al. (2009) and Ranade and Thiagarajan (2017). Furthermore, we refer the interested readers to Najmi et al. (2019b) where an application of the CCD model to calibrate an agent-based model is discussed.

**Figure C1:**
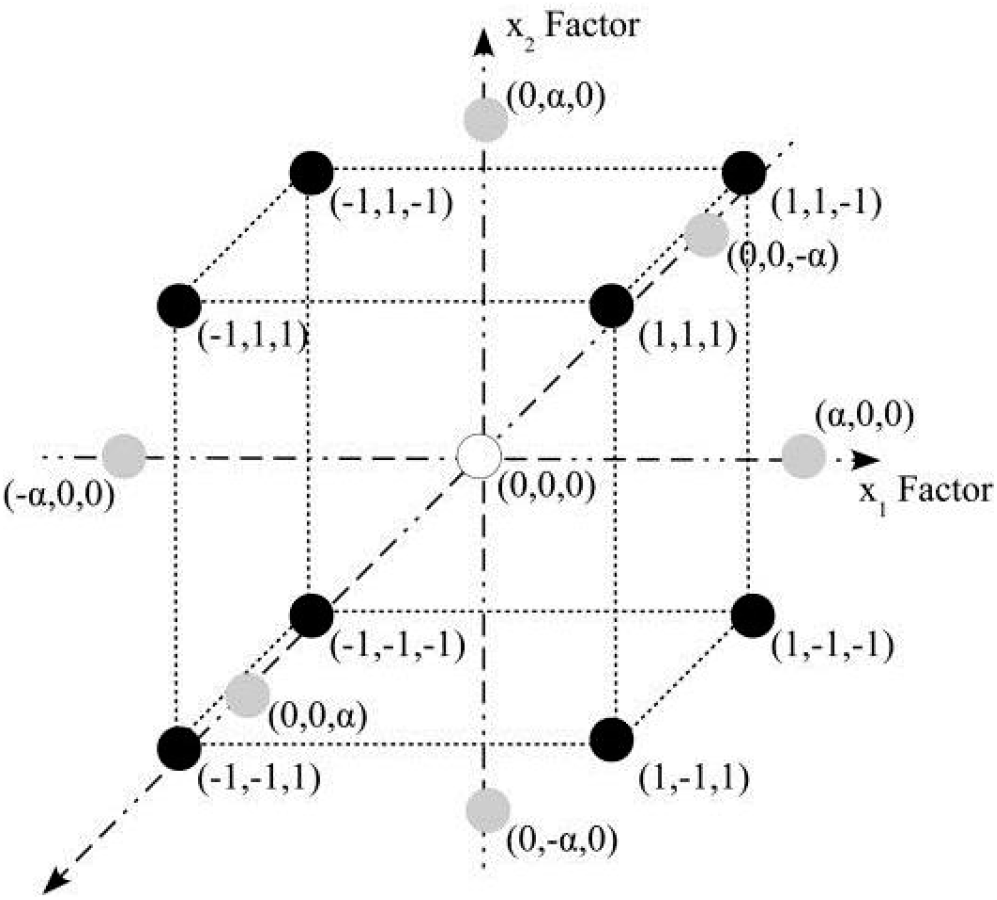
Schematic diagram of a three-factor central composite design (CCD)

Applied to the disease transmission model calibration, the factors are the parameters that should be considered for calibration and responses are the model outputs that should be reproduced. In the following paragraphs, we illustrate the parameters and model outputs that we have considered for calibration. Note that for each parameter, we define a feasible range from which its optimum value should be determined. A value is the best when its interaction with other parameters results in the best model performance (Najmi et al., 2019a). Not only may the values in a feasible range not have the same probability to be selected, but also having a uniform probability for the values may generate poor results. Thus, we use values that have already been estimated in other studies for some of the parameters and penalise deviations from these values.

Different incubation periods have been reported in various studies, including 2 days in Chang et al. (2020) and 5-6 days in Anderson et al. (2020). However, most of the researchers have reported the parameter to be around 4 days (Guan et al., 2020). So, we use 3 to 5 days for the feasible range of incubation period paramerter from which the values closer to 4 are considered to be more desirable and have a higher chance of being selected. We assume that the contact number for all activity types is a function of occupation and is within the range of 1 to 3. While there is no evidence concerning a generally applicable contact rate in the literature, as for incubation period parameter, we assign a higher chance (and so higher desirability) to the values close to 2. This is very helpful, as it reduces the degree of freedom in the calibration of parameters. We also assume that agents with sales (including sales workers) and general (including community and personal service workers, clerical and administrative workers) occupations have higher contact rates compared to the agents with other occupation types with a coefficient placed in the range 1 to 1.5.

Contact intensity in PT vehicles is a controversial parameter. Muller et al. (2020) assume that the intensity is 10 times higher in PT vehicles than in activities; so, we consider the feasible range of 6 to 14. While some of the cases are asymptomatic with mild disease, they can be traced and isolated if they have been in contact with a household member of colleague, who is symptomatic and already isolated (Ferguson et al., 2020).

So, we can assume all the infected cases have the same daily probability to be traced and quarantined with a daily probability within the range of 0.05 to 0.15. The reproduction number is another parameter that recently has been frequently reported, with most of the estimations placed between 3 and 5 (Liu et al., 2020; Anastassopoulou et al., 2020). Dividing the values by the number of infectious days, average contact number and average daily number of trips results in an estimation of the feasible range for infection probability per trip in the range of 0.03 to 0.05. Note that we do not differentiate between symptomatic and asymptomatic cases and assume that all the infected cases have the same infectious rates. Table C1 summaries the selected parameters and their feasible ranges.

**Table C1:**
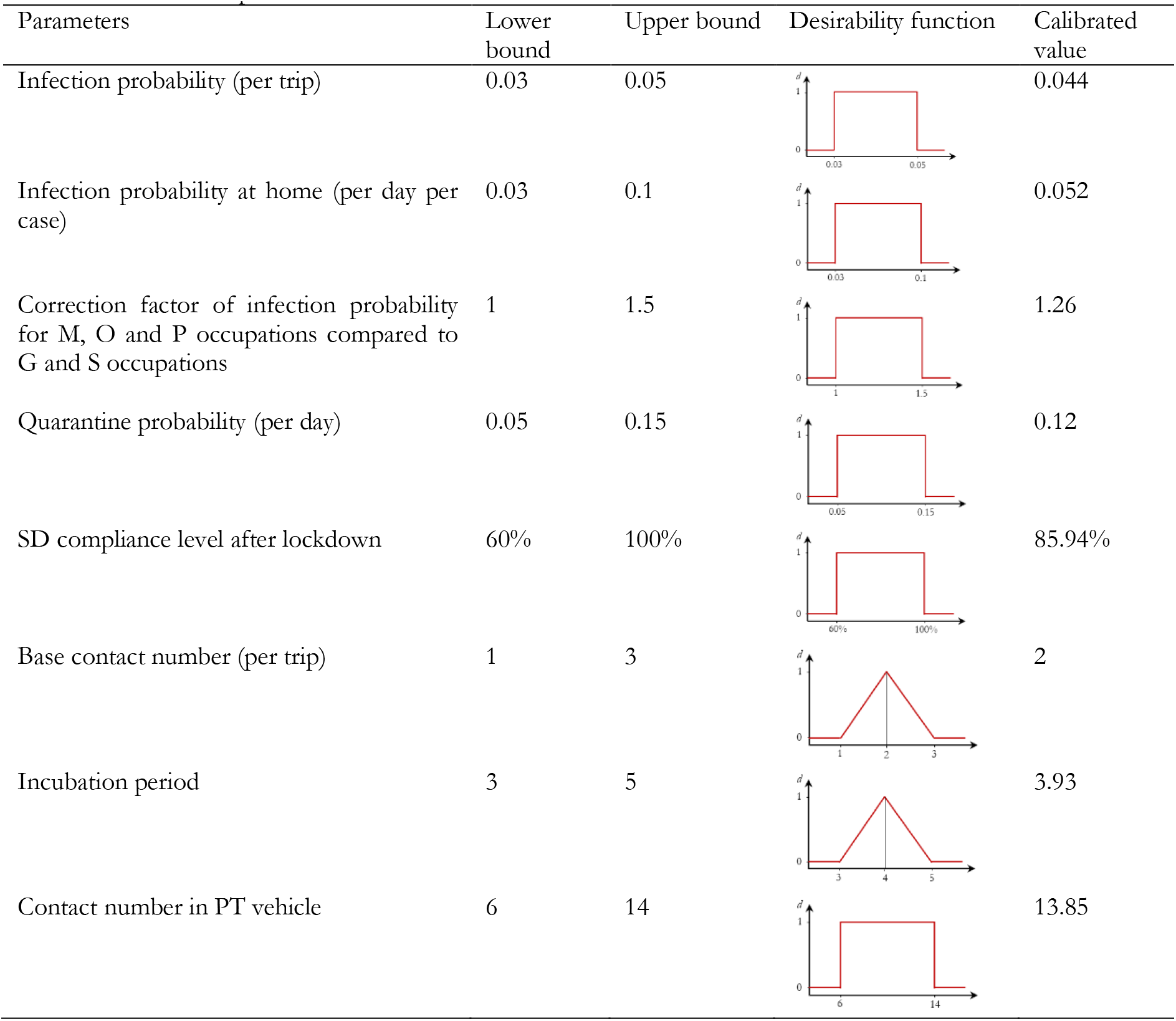
Calibrated parameters

| Parameters | Lower bound | Upper bound | Desirability function | Calibrated value |
| --- | --- | --- | --- | --- |
| Infection probability (per trip) | 0.03 | 0.05 |  | 0.044 |
| Infection probability at home (per day per case) | 0.03 | 0.1 |  | 0.052 |
| Correction factor of infection probability for M, O and P occupations compared to G and S occupations | 1 | 1.5 |  | 1.26 |
| Quarantine probability (per day) | 0.05 | 0.15 |  | 0.12 |
| SD compliance level after lockdown | 60% | 100% |  | 85.94% |
| Base contact number (per trip) | 1 | 3 |  | 2 |
| Incubation period | 3 | 5 |  | 3.93 |
| Contact number in PT vehicle | 6 | 14 |  | 13.85 |

The daily and cumulative infected cases are the best-defined measures of disease spread in urban areas, with reliable data generally being publicly available. Thus, we use maximise predicted fit to these two variables as the objective for parameter calibration. Furthermore, as in Muller et al. (2020), we assume that about a quarter of infections occur in PT vehicles, as the third objective to fit. Note that replication of death rate is another important objective to meet; however, the interaction of the death rate parameter on the model performance is not significant. Thus, the reproduction of number of death individuals can be easily met.

Once the model parameters and objective functions have been defined, experiments to explore the parameter solution spaced need to be conducted. The number of experiments and their design is a function of desired accuracy, and the number of selected parameters for calibration. For more information about experimental designs, readers are referred to Ahn (2015). After conducting the experiments and calculating their respective response values, the functional relationships in Eq. (1) can be obtained. The next step is to use the developed functional relationships in an optimisation formulation and find the optimum values for the parameters. In other words, we seek the best combination of parameters that can reproduce the observed statistics of daily number of cases, cumulative number of cases, and proportion of infections occurring in PT. We use simultaneous optimisation using desirability functions, as popularised by Derringer and Suich (1980). In this technique, each parameter x_i_ and response *y*_*j*_ is converted into a desirability function (*d*_*i*_ and *d*_*j*_ respectively) that varies between 0 and 1. If a parameter x_*i*_ is outside its feasible range, then *d*_*i*_ = 0, otherwise, it receives a desirability index based on a desirability function defined for the parameter. Based on the feasible ranges and their desirabilities that we defined before, the desirability functions are generated and shown in Table C1. The same definition applies for parameters and their desirability functions, except that the desirability functions are different than those for parameters. To quantify the power of the model to reprocude the observed data a function (index) is needed to measure the closeness of the simulated outputs to observed statistics. Root mean square error (RMSE) and absolute deviation are common functions used in the literature. The smaller the indexes are, the better the model fit. So, the target values are zero and values close to it have highest desirability (See Table C2). The design parameters are determined by solving the formulation in Eq. 2 which seeks the best combination of parameter values that maximise the overall desirability of the system (Myers et al. 2009).

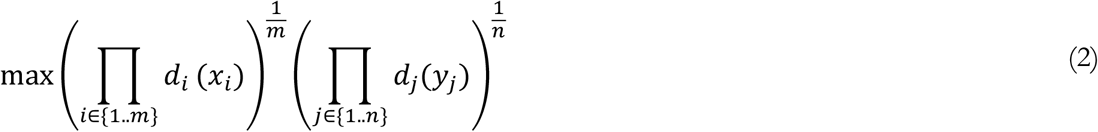

Subject to:

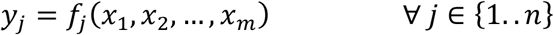

In this equation, *m* and *n* are the total number of parameters and the total number of objective criteria, respectively. The interested readers are referred to Najmi et al. (2019b) for detailed description about the calibration procedure. Using the simultaneous optimisation on the desirability functions the best values for the disease-specific parameters are obtained and given in the last column of Table C1. Most importantly, the calibrated value for SD compliance level after lockdown is 85.9% which means that the contact numbers have been reduced by 85.9% in Sydney GMA after lockdown.

**Table C2:**
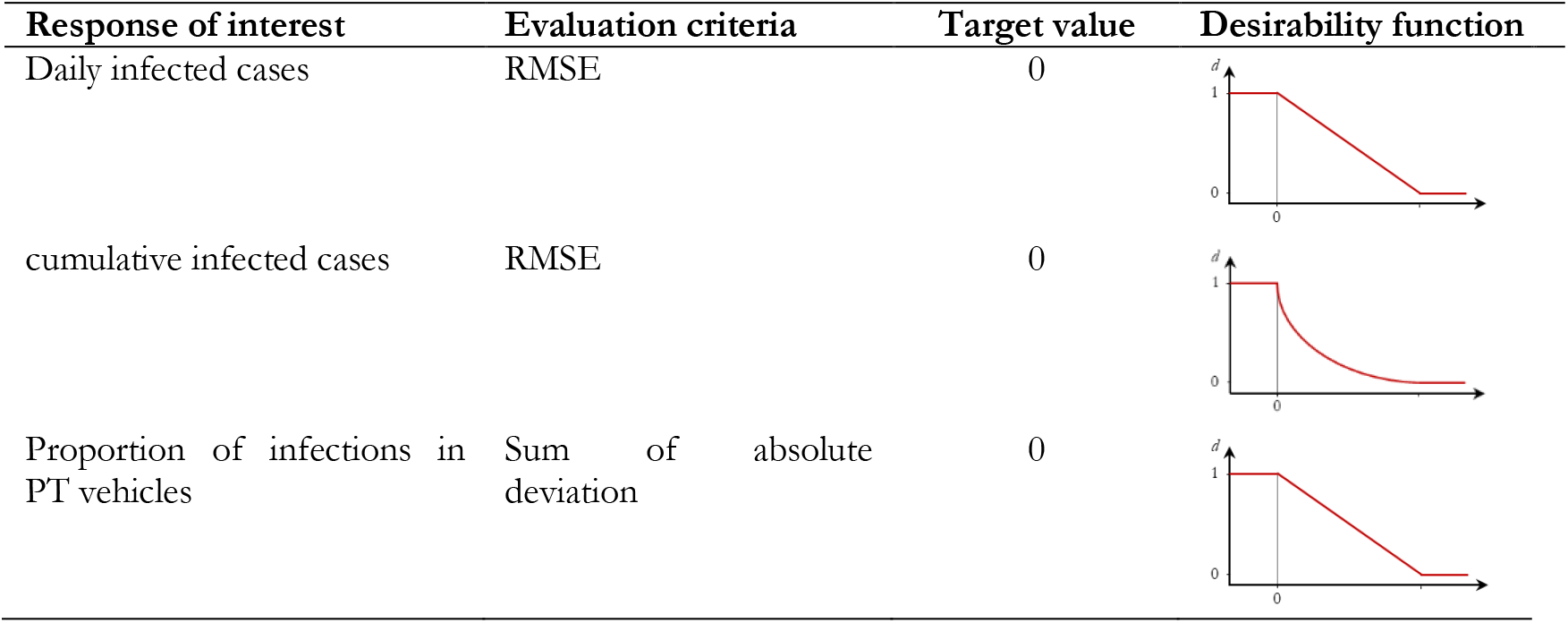
Objective criteria for calibration

| Response of interest | Evaluation criteria | Target value | Desirability function |
| --- | --- | --- | --- |
| Daily infected cases | RMSE | 0 |  |
| cumulative infected cases | RMSE | 0 |  |
| Proportion of infections in PT vehicles | Sum of absolute deviation | 0 |  |

